# Parental genetically predicted liability for coronary heart disease and risk of adverse pregnancy outcomes

**DOI:** 10.1101/2023.08.18.23294257

**Authors:** Álvaro Hernáez, Karoline H. Skåra, Christian M. Page, Vera R. Mitter, Marta H. Hernández, Per Magnus, Pål R. Njølstad, Ole A. Andreassen, Elizabeth C. Corfield, Alexandra Havdahl, Øyvind Næss, Ben Brumpton, Bjørn Olav Åsvold, Deborah A. Lawlor, Abigail Fraser, Maria Christine Magnus

## Abstract

**Background:** Adverse pregnancy outcomes (APO) may unmask a woman’s underlying risk for coronary heart disease (CHD). To test this, we estimated associations between genetically predicted liability for CHD and risk of APOs in mothers and their male partners. We hypothesized that associations would be found for women, but not their partners (negative controls).

**Methods:** We studied up to 83,969 women (and up to 55,568 male partners) participating in the Norwegian Mother, Father and Child Cohort Study or the Trøndelag Health Study with genotyping data and information on history of any APO in their pregnancies (miscarriage, stillbirth, hypertensive disorders of pregnancy, gestational diabetes, small for gestational age, large for gestational age, and spontaneous preterm birth). Maternal and paternal genetic risk scores (GRS) for CHD were generated using 148 gene variants (*p*-value < 5 × 10^-8^, not in linkage disequilibrium). Associations between GRS for CHD and each APO were determined using logistic regression, adjusting for genomic principal components, in each cohort separately, and combined using fixed effects meta-analysis.

**Results:** One standard deviation increase in the GRS for CHD in women was related to increased risk of any hypertensive disorders of pregnancy (odds ratio [OR] 1.08, 95% confidence interval [CI] 1.05-1.10), pre-eclampsia (OR 1.08, 95% CI 1.05-1.11), and small for gestational age (OR 1.04, 95% CI 1.01-1.06). Imprecise associations with lower odds of large for gestational age (OR 0.98, 95% CI 0.96 to 1.00) and higher odds of stillbirth (OR 1.04, 95% CI 0.98 to 1.11) were suggested. These findings remained consistent after adjusting for number of total pregnancies and the male partners’ GRS and restricting analyses to stable couples. Associations for miscarriage, gestational diabetes, and spontaneous preterm birth were close to the null. In male partners, there was weak evidence of an association with spontaneous preterm birth (OR 1.02 [0.99 to 1.05]), but not with other APOs.

**Conclusions:** Hypertensive disorders of pregnancy, small for gestational age and stillbirth unmask women with a genetically predicted existing propensity for CHD. The association of paternal genetically predicted CHD risk with spontaneous preterm birth needs further exploration.

## INTRODUCTION

Adverse pregnancy outcomes (APO), such as miscarriage, stillbirth, hypertensive disorders of pregnancy (HDP), gestational diabetes (GD), small for gestational age (SGA), large for gestational age (LGA), and spontaneous preterm birth (sPTB), are thought to unmask a woman’s underlying liability for cardiovascular disease, identifying women who ‘fail’ the cardiometabolic stress test of pregnancy [1]. Preconception cardiovascular risk factors are associated with APOs [2–7] and the extent of physiological changes of pregnancy including vasodilation, decreases in glucose, and changes in lipoprotein subclasses and biomarkers of low-grade inflammation [8]. Moreover, adjustment for preconception cardiovascular risk factors attenuates the relationship between APOs and maternal post-partum risk of cardiovascular disease [9–11]. An association between genetically predicted liability for cardiovascular disease and APOs in women would offer further support to the ‘unmasking’ hypothesis as germline variants are inherited at birth.

We therefore investigated associations between maternal and paternal genetic liability for coronary heart disease (CHD) with APOs (miscarriage, stillbirth, HDP, GD, SGA, LGA and sPTB). We hypothesized that we would find an association in women but not in men, as men’s genetic predisposition to CHD would not be revealed when their female partner undergoes the cardiometabolic stress test of pregnancy. Hence, we considered men as imperfect negative controls. Imperfect, because shared family environment associated with both genetic liability for CHD and APO risk (e.g., body mass index, smoking) [12] may result in associations between genetic liability for CHD and APO risk in men. Paternal genetic variants linked to CHD might also impact the epigenetics of their reproductive cells or the quality of their sperm quality, and these factors may in turn influence APO risk [13, 14]. Finally, fetal genetics inherited from both parents may also affect risk of APOs [15, 16]. We used genetically predicted CHD risk as the exposure because CHD is the most common cardiovascular disease, the leading cause of death globally, and the cardiovascular event with the highest proportion of genetic variance explained in genome-wide association studies (GWAS) [17].

## METHODS

### Population description

We studied participants in the Norwegian Mother, Father, and Child Cohort Study (MoBa) [18] and the Trøndelag Health Study (HUNT) [19]. MoBa is a pregnancy cohort study led by the Norwegian Institute of Public Health in which pregnant women and their partners were recruited at approximately 17 gestational weeks between 1999 and 2008 all over Norway. The participation rate was 41%, and the cohort includes approximately 95,200 women and 75,200 of their male partners [18]. The HUNT study is a population-based cohort of the Trøndelag County in Norway (representative of the general adult Norwegian population regarding morbidity, mortality, income, and age), led by the Norwegian University of Science and Technology and based on four data collection surveys between 1984 and 2019 [19, 20]. Both MoBa and HUNT participants are of predominantly European ancestry.

Information from the Medical Birth Registry of Norway (MBRN) for participants in both cohorts was obtained by linkage using unique identification numbers. This work is based on a subsample of participants in the two cohorts who had at least one registered singleton pregnancy in the MBRN, available genotype data, and information on the pregnancy outcomes of interest (**Figure 1**). Regarding HUNT, as the MBRN has information on all births in Norway from 1967 onwards [21], we restricted our analyses to cohort participants who were 15 years or younger at the time when the MBRN was set up (born 1952 or later), to capture all of their births. Genotype data in both cohorts came from biological samples obtained from participants after genotype calling, imputation, and quality control [20, 22]. This work is presented according to the Strengthening the Reporting of Observational Studies in Epidemiology guidelines.

**Figure 1.**
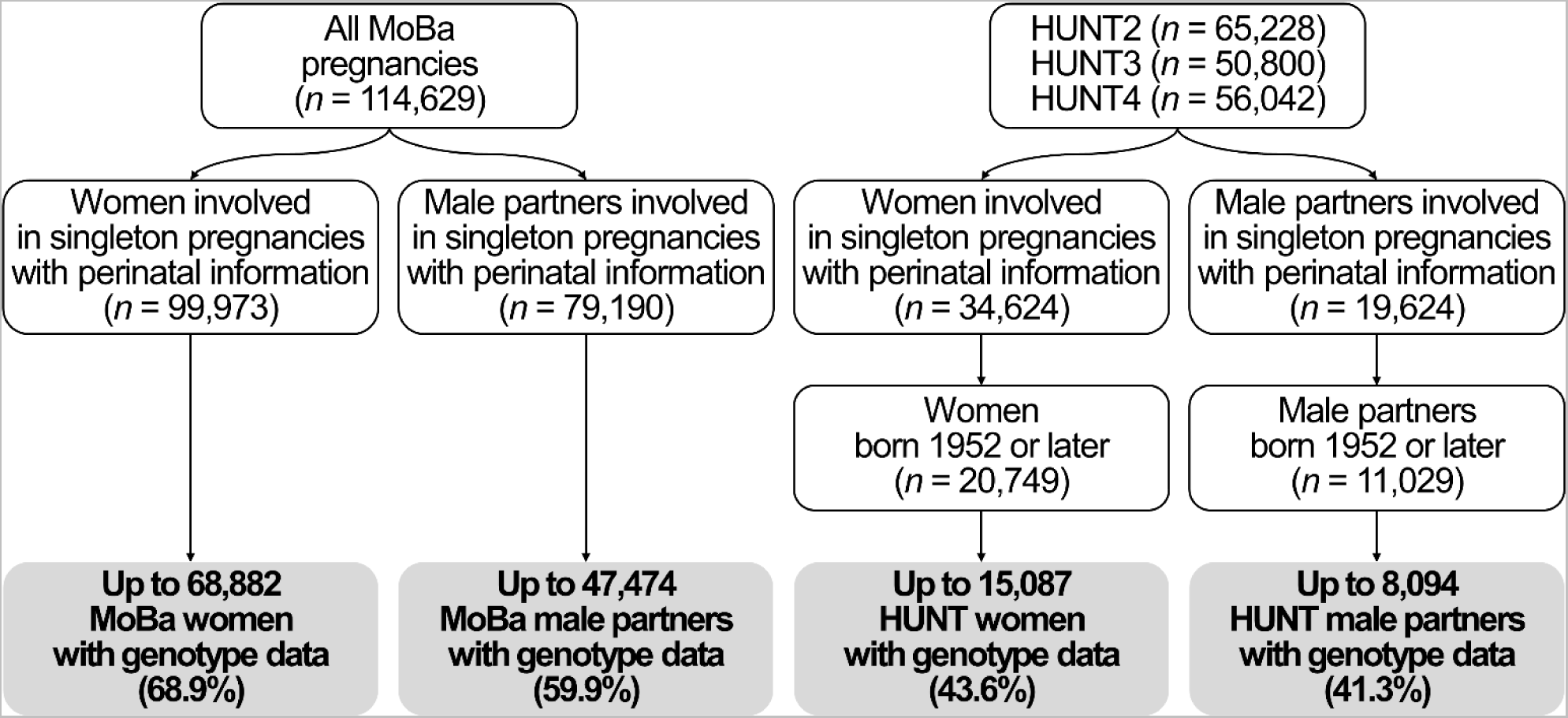
Flow chart

### Genetic liability for coronary heart disease

We obtained publicly available summary data on the genetic variants linked to CHD from the latest GWAS including ∼550,000 individuals from the UK Biobank and CARDIoGRAMplusC4D cohorts [17]. No MoBa or HUNT participants took part in this GWAS. It identified 148 genetic variants (single nuclear polymorphisms) associated with CHD at genome-wide significance (*p*-value < 5 × 10^-8^) and independent of each other (i.e., not in linkage disequilibrium [pairwise *r*^2^ <0.1 and a physical distance of at least 5,000bps]) [23]. 141 and 146 of these genetic variants were available in the MoBa and HUNT databases, respectively. We used these variants to calculate weighted genetic risk scores (GRSs) for CHD, representing the genetic liability for CHD [24]. The exact genetic variants used in each cohort is available in **Supplemental Table 1**.

### Adverse pregnancy outcomes

APOs were defined using information from the MBRN for both cohorts. We identified individuals in MoBa and HUNT with any history of any APO across all their registered pregnancies from 1967 to 2019. Miscarriage was defined as any fetal loss prior to 23 completed gestational weeks, while stillbirth was defined as any fetal death after 23 completed gestational weeks, as registered in the MBRN [21]. The MBRN contains self-reported information on the number of prior miscarriages and stillbirths at the time of each registered pregnancy (it is not possible to distinguish the exact gestational age of prior pregnancies not registered in the MBRN), in addition to the status and gestational age of the registered pregnancies. We combined the self-reported information on previous pregnancies and the registered pregnancies to define complete history of miscarriage and stillbirth. HDP was defined as having any registration of gestational hypertension, preeclampsia, eclampsia, or Hemolysis, Elevated Liver enzymes and Low Platelets syndrome in the absence of hypertension prior to the pregnancy [25]. Registrations of GD in the MBRN are made according to the criteria of the Norwegian Society of Gynecology and Obstetrics in the absence of a history of diabetes [26]. Notably, there is no information on blood pressure measurements, proteinuria levels or glucose levels in the MBRN. The registrations of HDP and GD are therefore made based on the national clinical guidelines at the time [25, 26]. Gestational age was estimated using ultrasound around 18 completed gestational week or last menstrual period for the small proportion without an ultrasound assessment. SGA was defined as birth weight <10^th^ percentile for gestational age and biological sex [27], whilst LGA was defined as birth weight ≥90^th^ percentile for gestational age and biological sex of the offspring. Participants who delivered an SGA baby were excluded from the analyses on LGA and vice versa. Any non-medically induced delivery prior to the 37^th^ week of pregnancy was considered a sPTB (induced deliveries and C-sections were excluded from the reference group) [28].

### Descriptive variables

To describe the characteristics of our population, we obtained information on birth year (value in the first pregnancy in MBRN, continuous), the number of pregnancies in which the participants were involved (continuous), whether the study participants were involved in pregnancies with more than one partner (yes/no), years of education (value registered in the first MoBa/HUNT questionnaire available, continuous), body mass index (value registered in the first MoBa/HUNT questionnaire available, in kg/m^2^), having ever smoked (yes/no), and highest obtained parity (stillbirths plus live births according to the MBRN, continuous) [29, 30].

### Ethical approval and consent to participate

Both MoBa and HUNT follow the Declaration of Helsinki for Medical Research on human subjects. MoBa is currently regulated by the Norwegian Health Registry Act and its data collection is approved by the Norwegian Data Protection Authority.HUNT was authorized by the Regional Committee for Medical and Health Research (Health Region IV, Norway). Participants provided a written informed consent before joining the cohorts. This project was particularly approved by the Regional Committee for Medical and Health Research Ethics of South/East Norway (references: 2017/1362 and 78545).

### Statistical analyses

We describe the distributions of normally distributed continuous variables using means and standard deviations (SDs), non-normally distributed continuous variables using medians and 1^st^-3^rd^ quartiles, and categorical variables using numbers and percentages. We assessed differences between participants with and without genotype information using t-tests (normally distributed continuous variables), Mann-Whitney U-tests (non-normally distributed continuous variables), and chi-squared tests (categorical variables).

We studied the association between a one SD higher GRS for CHD and risk of APOs, in mothers and their male partners separately, using logistic regression. We analyzed MoBa and HUNT data individually, and subsequently meta-analyzed their estimates using a fixed effects model, assuming participants from both cohorts come from the same underlying population and any difference in associations is random. We tested this assumption using the Cochrane Q-test for between-study heterogeneity, using a *p*-value threshold of <0.1 as an indication of heterogeneity given the statistical inefficiency of between-study heterogeneity tests and the smaller sample size of HUNT compared to MoBa. If significant heterogeneity was detected for any of the outcomes, we meta-analyzed the estimates of the two studies using a random effects model as a complementary analysis. We reduced confounding of the relationship between GRSs for CHD and the outcomes due to population stratification by adjusting our analyses for the first 20 ancestry-informative principal components [31]. Analyses were further adjusted for genotype batch.

As parental negative control analyses (i.e., comparing maternal associations to paternal associations) can be biased by assortative mating and other shared if they are not mutually adjusted for each other [32], we repeated our main analyses in the subgroup of mothers and their partners where both had genetic data. We present results in this subgroup without and with mutual adjustment. The former analysis, when compared with the main results, explores evidence of selection bias in the subsample in couples with genetic data.

We conducted additional sensitivity analyses. Firstly, the more pregnancies a person has, the higher the likelihood of detecting an effect of parental genetic predisposition to CHD on the risk of APO. To minimize this bias, we further adjusted for the total number of pregnancies the participant had. Secondly, a participant who had pregnancies with various partners might have a differing risk of APO with each partner. Therefore, we performed another sensitivity analysis that was confined to stable couples only (i.e., participants without pregnancies with different partners according to the MBRN).

## Software

We conducted our analyses in R Software v. 4.0.3. Our analysis code is available in https://github.com/alvarohernaez/GRS_CHD_pregnancy_complications_MoBa_HUNT/.

## RESULTS

### Study population

Our analyses were conducted in up to 68,882 women and 47,474 of their male partners in MoBa, and up to 15,087 women and 8,094 of their male partners in HUNT (**Figure 1**), described in **Table 1**. Meta-analyses of the main analyses involved between 66,110 (SGA) and 83,900 women (stillbirth and GD) and between 43,967 (SGA) and 55,790 men (stillbirth and GD). The exact sample sizes for all analyses are available in **Supplementary Table 2**.

**Table 1.** Population description.

|  | MoBa |  | HUNT |  |
| --- | --- | --- | --- | --- |
|  | Women<br>(*n* = 68,882) | Male partners<br>(*n* = 47,474) | Women<br>(*n* = 15,087) | Male partners<br>(*n* = 8,094) |
| Birth year, range | 1954-1993 | 1938-1990 | 1952-1988 | 1952-1988 |
| Number of pregnancies,<br>median (1<sup>st</sup>-3<sup>rd</sup> quartile; range) | 2 (2-3)<br>[range: 1-14] | 2 (2-3)<br>[range: 1-14] | 2 (2-3)<br>[range: 1-10] | 2 (2-3)<br>[range: 1-10] |
| Participants in pregnancies<br>with more than one different<br>partner, *n* (%) | 9,147<br>(13.3%) | 5,819<br>(12.3%) | 745<br>(4.74%) | 178<br>(2.08%) |
| Age in first pregnancy, median<br>(1<sup>st</sup>-3<sup>rd</sup> quartile) | 27<br>(24-30) | 29<br>(26-33) | 24<br>(20-27) | 26<br>(23-29) |
| Education years,<br>mean ± SD | 17.0 ± 3.36 | 16.3 ± 3.56 | 14.3 ± 4.13 | 13.5 ± 4.08 |
| Body mass index (kg/m<sup>2</sup>), | 23.1 | 25.4 | 25.2 | 26.6 |
| median (1<sup>st</sup>-3<sup>rd</sup> quartile) | (21.1-25.9) | (23.6-27.7) | (22.7-28.6) | (24.5-29.1) |
| Ever smokers, *n* (%) | 35,631<br>(52.3%) | 24,294<br>(51.2%) | 8,865<br>(57.0%) | 4,233<br>(50.1%) |
| Total number of deliveries: |  |  |  |  |
| 0, *n* (%) | 0<br>(0%) | 0<br>(0%) | 1,598<br>(10.5%) | 595<br>(7.00%) |
| 1, *n* (%) | 4,611<br>(6.71%) | 2,894<br>(6.12%) | 6,849<br>(45.1%) | 4,041<br>(47.5%) |
| 2, *n* (%) | 32,186<br>(46.8%) | 23,368<br>(49.4%) | 5,088<br>(33.5%) | 3,056<br>(36.0%) |
| 3, *n* (%) | 24,121<br>(35.1%) | 16,357<br>(34.6%) | 1,285<br>(8.46%) | 654<br>(7.69%) |
| 4 or more, *n* (%) | 7,784<br>(11.3%) | 4,671<br>(9.88%) | 378<br>(2.49%) | 154<br>(1.81%) |
| Miscarriage,<br>*n* (%) | 20,866<br>(30.4%) | 14,451<br>(30.6%) | 1,950<br>(30.0%)<sup>a</sup> | 1,145<br>(29.6%)<sup>a</sup> |
| Stillbirth,<br>*n* (%) | 872<br>(1.27%) | 581<br>(1.23%) | 198<br>(1.30%) | 84<br>(0.99%) |
| Hypertensive disorders of<br>pregnancy, *n* (%) | 7,288<br>(10.6%) | 5,186<br>(11.0%) | 1,500<br>(9.87%) | 870<br>(10.2%) |
| Pre-eclampsia + eclampsia, *n*<br>(%) | 5,143<br>(7.49%) | 3,589<br>(7.59%) | 1,080<br>(7.11%) | 641<br>(7.54%) |
| Gestational diabetes,<br>*n* (%) | 1,495<br>(2.18%) | 1,041<br>(2.20%) | 127<br>(0.84%) | 62<br>(0.73%) |
| Small for gestational age,<br>*n* (%) | 10,552<br>(15.8%) | 7,354<br>(16.0%) | 1,544<br>(10.2%) | 862<br>(10.1%) |
| Large for gestational age,<br>*n* (%) | 14,620<br>(21.7%) | 9,880<br>(21.3%) | 1,733<br>(11.4%) | 1,070<br>(12.6%) |
| Spontaneous preterm birth,<br>*n* (%) | 6,282<br>(9.18%) | 4,187<br>(8.89%) | 2,643<br>(17.5%) | 1,509<br>(17.8%) |

<sup>a</sup>: analyses restricted to 1998 or later pregnancies.

MoBa participants with genotype information were not meaningfully different in their birth year, total number of pregnancies, proportion of stable couples, age at first pregnancy, years of education, body mass index, smoking, or number of deliveries in relation to those without genotype data. However, they were slightly less likely to have experienced stillbirth, GD, SGA, and sPTB (**Supplementary Table 3**). HUNT participants with genotype data were born on average 12 years earlier but were not meaningfully different on their total number of pregnancies, proportion of stable couples, age at first birth, years of education, body mass index, and smoking. However, they were more like to experience several APOs (stillbirth, HDP, SGA, LGA, and sPTB) and less likely to have a history of GD (**Supplemental Table 3**).

### Genetically predicted CHD and adverse pregnancy outcomes

**Figure 2** shows the main analyses and all additional and sensitivity analyses for women (Panel A) and men (Panel B).

**Figure 2.**
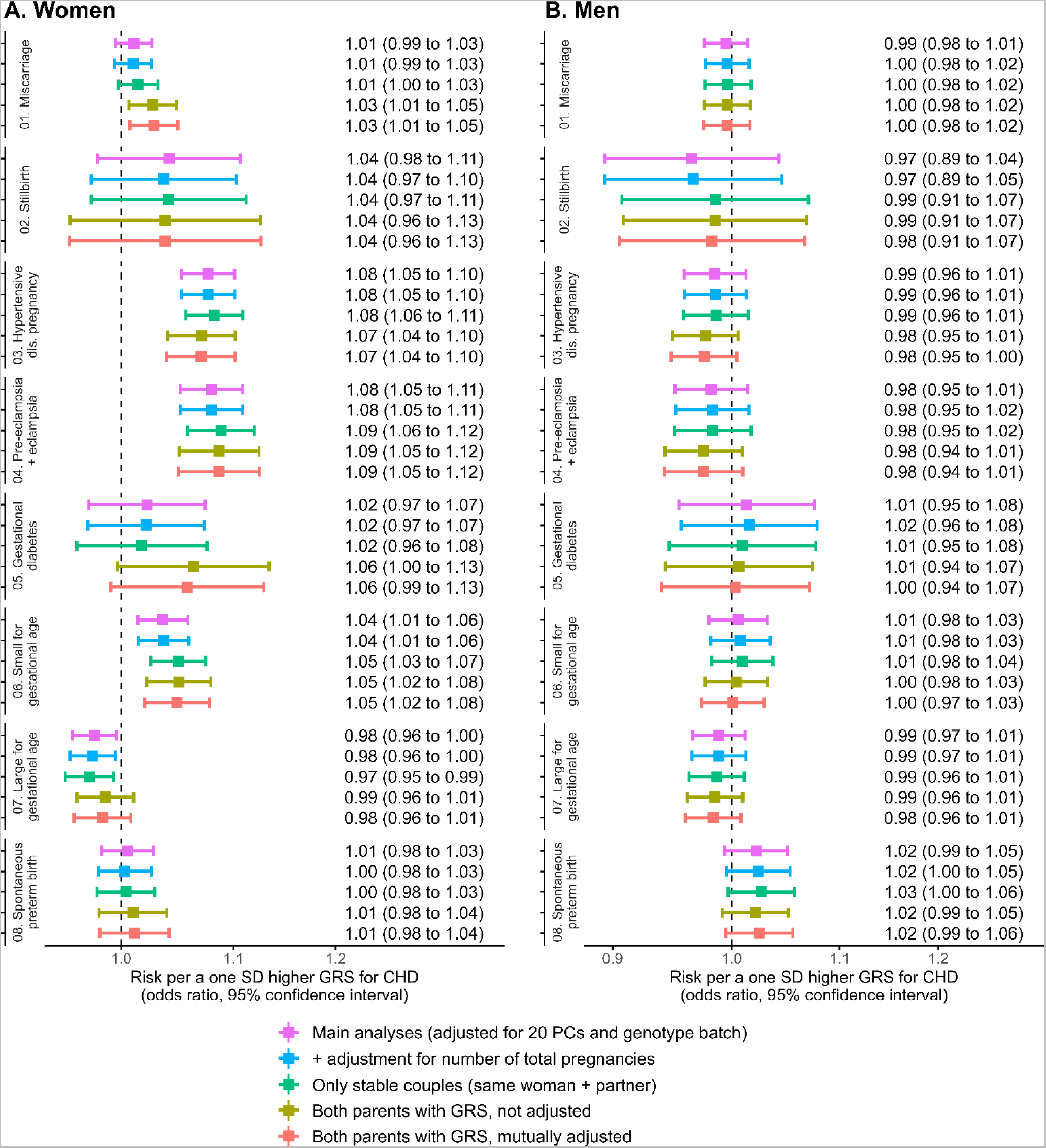
Associations between a one SD higher genetic risk score for coronary heart disease and pregnancy outcomes in women (A) and men (B).

In women, a one SD higher GRS for CHD was associated with 8% greater odds of HDP (OR 1.08, 95% CI 1.05 to 1.10), 8% greater odds of preeclampsia (OR 1.08, 95% CI 1.05 to 1.11), and 4% greater odds of having an SGA baby (OR 1.04, 95% CI 1.01 to 1.06). There was evidence of an inverse association between the GRS for CHD and LGA (+1 SD in GRS: OR 0.98, 95% CI 0.96 to 1.00). A positive but imprecise relationship was found for stillbirth (OR 1.04, 95% CI 0.98 to 1.11) and no association was found with sPTB. These findings remained consistent in the subset with genetic data on both parents when adjusting for partners’ GRS, as well as in sensitivity analyses adjusting for the number of total pregnancies and restricting the analyses to stable couples. Stronger associations between the GRS for CHD and the risk of miscarriage and GD were observed in couples with genotype information in both parents.

In male partners, no association between the GRS for CHD and any APO was observed, except for evidence of a relationship between the GRS for CHD and greater odds of sPTB (OR 1.02, 95% CI 0.99 to 1.05). Estimates remained robust in all sensitivity analyses.

Between-study heterogeneity was detected for two outcomes: the association of the GRS for CHD with GD in women (MoBa: OR 1.04, 95% CI 0.99 to 1.09; HUNT: OR 0.86, 95% CI 0.72 to 1.03; *p*-value for between-study heterogeneity = 0.045; random effects meta-analysis: OR 0.96, 95% CI 0.30 to 3.09), and with sPTB in men (MoBa: OR 1.04, 95% CI 1.01 to 1.07; HUNT: OR 0.97, 95% CI 0.92 to 1.03; *p*-value for between-study heterogeneity = 0.053; random effects meta-analysis: OR 1.01, 95% CI 0.68 to 1.50). For the rest of the outcomes, findings were consistent between the two cohorts (**Supplemental Figure 1**).

Sample sizes vary from 40,703 to 83,900 in women, and from 39,334 to 55,790 in male partners, being smallest for the analyses in the subgroup with genetic data on both parents (see Supplementary Table 2 for information on sample sizes).

## DISCUSSION

Maternal genetic CHD risk was associated with increased risk of HDP, preeclampsia, and SGA. Weak evidence with confidence intervals spanning the null was found for associations with greater risk of stillbirth and reduced risk of LGA. Null associations with miscarriage, GD, and sPTB were observed. We also found weak evidence of an association between genetic CHD risk with sPTB in men, with associations with other outcomes in men all being close to the null.

Our findings support the hypothesis that a woman’s existing predisposition to cardiovascular disease may be revealed during pregnancy, specifically through HDP and SGA [1]. The lack of association between male partners’ GRS for CHD and these conditions supports these APOs reflecting maternal pre-existing risk of cardiovascular disease. This agrees with previous evidence showing that women who developed these APOs had a greater burden of cardiovascular risk factors before pregnancy [5, 6]. Moreover, high levels of genetically determined cardiovascular risk factors, such as blood pressure, body mass index, and type-2 diabetes, have been linked to a higher likelihood of preeclampsia or eclampsia [33]. Finally, a recent maternal GWAS of preeclampsia and gestational hypertension found similar associations between the preeclampsia and gestational hypertension polygenic risk scores and hypertension and related outcomes as those seen in GWASs of men and non-pregnant women, leading authors to conclude that the loci associated with preeclampsia and gestational hypertension were not pregnancy specific, rather that pregnancy unmasks existing risk [34].

In relation to the association between the maternal genetically predicted CHD risk and other APOs, pregnancy loss is a heterogeneous condition, and maternal predisposition to cardiovascular disease may only be relevant to specific subsets of losses. Stillbirth, which was suggestively linked to greater genetic liability for CHD in our study, is thought to be induced by placental insufficiency and fetal growth restrictions due to some cardiovascular risk factors during pregnancy, such as hypertension [35–37]. Our findings also revealed an unexpected inverse association between the GRS for CHD and LGA. However, given the small magnitude of the relationship in comparison to the other APOs, and the established role of glucose metabolic disturbances (such as type 1 diabetes, GD or prediabetic states due to excess weight) in increasing the risk of having LGA babies [38–40], it would be advisable to validate the association between the GRS for CHD and LGA in future studies involving other populations. As for GD, it has been shown to be closely related to type 2 diabetes, including with a recent GWAS showing strong genetic correlation between the two [41]. Given the associations of type 2 and GD with cardiovascular disease [42, 43], it is surprising that we did not see an association of the maternal GRS for CHD with GD in our study. Limited cases in our population (∼2%), the between-study heterogeneity in MoBa and HUNT for this outcome, changes in diagnostic criteria for GD in Norway (which were large and occurred within the time frame of both studies) [26], and underrepresentation in the early years of the MBRN (the precise definition of GD, which differentiates between the diabetic status of the pregnant mother before and during pregnancy, was introduced in the MBRN registration forms in December 1998) [44] may contribute to systematic errors in its diagnosis and hinder our ability to find robust associations. Finally, the lack of association between maternal genetically predicted CHD and sPTB may be due to the fact that not all risk factors for this condition are cardiovascular-related [45].

We conceptualized paternal genetic CHD risk as an imperfect negative control, as an association between the paternal GRS for CHD and APO risk could not arise from a direct stress test of pregnancy but associations might arise from shared family environment, paternally transmitted genetic variants of the fetus [46] or a maternal immunological response to pregnancy [47]. The mutual adjustment of paternal for maternal GRS should limit the effect of shared family environment, making this an unlikely explanation for the association between the GRS for CHD in male partners and risk of sPTB. There is evidence of both maternal and paternal genetic effects on gestational duration [48]. However, this is not consistent with our findings (some evidence of an association in men and no strong evidence of such a relationship in women). Given the number of associations that we have explored, and the different association observed in MoBa and HUNT (affected by between-study heterogeneity), it is possible that the association of paternal GRS for CHD with sPTB is a chance finding, which we would suggest treating with caution unless replicated in other independent studies. Of note, the relationships between the maternal GRS for CHD and HDP, pre-eclampsia, SGA and stillbirth were not attenuated after adjusting for the male partners’ GRS for CHD, suggesting that these relationships are specific to women and supporting these APOs likely reflecting pre-existing cardiovascular risk.

Our study has some limitations. Firstly, male partners were considered as imperfect negative controls in our study, mainly due to the shared family environment that could lead to associations between paternal GRS for CHD and APOs. However, the mutual adjustment of maternal and paternal exposures in negative control analyses aims to control for these shared factors, including those occurring by assortative mating [32]. This imperfection may also be due to the fact that paternal genetic variants related to CHD could affect their reproductive cells’ epigenetics and quality and may impact certain placental characteristics that depend on the fetal genotype, which might subsequently alter the risk of APOs by alternative mechanisms [13–16]. Secondly, we identified differences between MoBa and HUNT participants with and without genotype data, which could result in selection bias and reduce our external validity. Differences in MoBa might stem from blood sampling during pregnancy or at delivery and it could be less likely if APOs had been detected. In HUNT, genotyped participants were older. However, selection bias had minimal impact on pregnancy outcomes when comparing MoBa with the general Norwegian population [49]. Thirdly, GD diagnosis may have been affected by systematic errors that may decrease our capacity to find robust associations with this outcome [26, 44]. Fourthly, we could not calculate sex-specific GRSs for CHD exposures due to the lack of sex-specific results in the largest GWAS on CHD. We assumed no sex differences in the GRS for CHD, but if untrue, our findings might be biased. Lastly, our study population’s characteristics (predominantly adult European men and women) limit the generalizability of our findings to other populations. Despite these limitations, our study has several strengths. To our knowledge, it is the first to investigate the role of genetic liability for CHD in APOs, conducted in a large, well- characterized, and genetically homogeneous population. This complements previous studies that have shown associations of cardiovascular risk factors (e.g., high blood pressure, dyslipidemia, impaired glucose tolerance) with APOs [2–7], by using a GRS that captures environmental (i.e., genetic risk of higher body mass, smoking, education, etc.) as well as likely biological risk for CHD. Unlike those previous studies, interpretation of the magnitude of a one SD difference in a genetic risk score for CHD is unclear. For example, the relative 8% increased odds for HDP and pre- eclampsia (OR 1.08, for both) appears small but must be understood in the context that the genetic variants that we used in the GRS explain ∼15% of the variation in CHD risk. We interpret our results as a qualitative indication of the association between a propensity to CHD influencing the risk of APOs, rather than trying to quantify that relationship.

## CONCLUSIONS

Our results indicate that pregnancy may unmask a woman’s pre-existing cardiovascular risk and suggest that a family history of cardiovascular disease or early onset of cardiovascular outcomes could help identify individuals who might benefit from advice and closer monitoring before and during pregnancy.

## DECLARATIONS

### Consent for publication

Not applicable.

### Availability of data and materials

The consent given by the MoBa and HUNT participants does not allow for storage of individual data in repositories or journals. Researchers who want to access MoBa datasets for replication should apply by sending an e-mail to. This procedure requires approval from the Regional Committee for Medical and Health Research Ethics in Norway and an agreement with MoBa. Data from the HUNT Study used for research are available upon request to the HUNT Data Access Committee to research groups who meet the data availability requirements described in http://www.ntnu.edu/hunt/data. Source data of the GWAS on CHD are available in the Online Table VI of the article of Van Der Harst P et al., Circ Res, 2018, available in the Supplemental Materials (https://www.ahajournals.org/doi/suppl/10.1161/CIRCRESAHA.117.312086), as well as in the IEU Open GWAS website (code: ebi-a-GCST005195).

### Author contributions

A.F. and M.C.M. conceived and designed the study, obtained funding, and coordinated the project. A.Hernaez, and M.C.M. are responsible for the data curation and the formal analysis. K.H.S. and C.M.P. provided support in data analysis, software use, and visualization of results. A.Hernaez, K.H.S., C.M.P., V.R.M., M.H.H., P.M., P.R.N, O.A., E.C., A.Havdahl, Ø.N., B.B., B.O.Å., D.A.L., A.F., and M.C.M. were involved in the definition of the methodology of the study and the interpretation of data. A.Hernaez prepared the first draft of the manuscript, and K.H.S., C.M.P., V.R.M., M.H.H., P.M., P.R.N, O.A., E.C., A.Havdahl, Ø.N., B.B., B.O.Å., D.A.L., A.F., and M.C.M. revised it critically.

## Acknowledgements

The MoBa Cohort Study is supported by the Norwegian Ministry of Health and Care Services and the Ministry of Education and Research. The HUNT Study is a collaboration between HUNT Research Centre (Faculty of Medicine and Health Sciences, Norwegian University of Science and Technology), Trøndelag County Council, Central Norway Regional Health Authority, and the Norwegian Institute of Public Health. The genotyping in HUNT was financed by the National Institutes of Health; University of Michigan; the Research Council of Norway; the Liaison Committee for Education, Research and Innovation in Central Norway; and the Joint Research Committee between St. Olavs Hospital and the Faculty of Medicine and Health Sciences, NTNU. We are grateful to all the participants in Norway who take part in the MoBa and HUNT studies, those who contributed to the recruitment, and the infrastructure surrounding both cohorts.

We thank the Norwegian Institute of Public Health for generating high-quality genomic data in the MoBa study. This research is part of the HARVEST collaboration, supported by the Research Council of Norway (#229624). We also thank the NORMENT Centre for providing genotype data, funded by the Research Council of Norway (#223273), South East Norway Health Authority, and Stiftelsen Kristian Gerhard Jebsen. We further thank the Center for Diabetes Research (University of Bergen) for providing genotype information and performing quality control and imputation of the data, in the context of research projects funded by the European Research Council Advanced Grant SELECTionPREDISPOSED, Stiftelsen Kristian Gerhard Jebsen, the Trond Mohn Foundation, the Research Council of Norway, the Novo Nordisk Foundation, the University of Bergen, and the Western Norway Health Authority.

This work was developed on the TSD (Tjeneste for Sensitive Data) online facilities, owned by the University of Oslo, which are operated and developed by the TSD service group at the IT-Department in University of Oslo. This paper does not necessarily reflect the position or policy of the Norwegian Research Council.

## List of abbreviations

APO: adverse pregnancy outcome
CHD: coronary heart disease
CI: confidence interval
GD: gestational diabetes
GRS: genetic risk score
GWAS: genome-wide association study
HDP: hypertensive disorders of pregnancy
HUNT: Trøndelag Health Study
LGA: large for gestational age
MBRN: Medical Birth Registry of Norway
MoBa: Mother, Father, and Child Cohort Study
OR: odds ratio
SGA: small for gestational age
SD: standard deviation
sPTB: spontaneous preterm birth

## SUPPLEMENTAL TABLES

**Supplemental Table 1.** Genetic variants used in the calculation of the genetic risk score for coronary heart disease.

| RSID | Chromosome | Position | Reference allele | Effect allele | Effect allele frequency | Beta | Standard error | p-value | Available in MoBa | Available in HUNT |
| --- | --- | --- | --- | --- | --- | --- | --- | --- | --- | --- |
| rs36096196 | 1 | 2252205 | C | T | 0.1483 | 0.0469 | 0.0082 | 1.33E-08 | Yes | Yes |
| rs2493298 | 1 | 3325912 | C | A | 0.1355 | 0.0514 | 0.0084 | 9.97E-10 | Yes | Yes |
| rs4072980 | 1 | 38456106 | G | A | 0.4303 | -0.0326 | 0.0052 | 4.13E-10 | Yes | Yes |
| rs11591147 | 1 | 55505647 | G | T | 0.0164 | -0.2406 | 0.0247 | 1.86E-22 | No | Yes |
| rs17114046 | 1 | 56966350 | G | A | 0.9062 | 0.096 | 0.0088 | 8.04E-28 | Yes | Yes |
| rs602633 | 1 | 109821511 | G | T | 0.2343 | -0.0999 | 0.0062 | 3.63E-58 | Yes | Yes |
| rs11806316 | 1 | 115753482 | G | A | 0.3668 | -0.0337 | 0.0054 | 4.16E-10 | Yes | Yes |
| rs11204892 | 1 | 151768048 | G | A | 0.1892 | -0.0487 | 0.0073 | 2.85E-11 | Yes | Yes |
| rs4845625 | 1 | 154422067 | C | T | 0.4338 | 0.0395 | 0.0051 | 6.69E-15 | Yes | Yes |
| rs1892094 | 1 | 169094459 | C | T | 0.4956 | -0.0362 | 0.0052 | 3.14E-12 | Yes | Yes |
| rs6700559 | 1 | 200646073 | C | T | 0.4715 | -0.0284 | 0.005 | 1.77E-08 | Yes | Yes |
| rs2820315 | 1 | 201872264 | C | T | 0.3012 | 0.0359 | 0.0056 | 1.10E-10 | Yes | Yes |
| rs60154123 | 1 | 210468999 | C | T | 0.1501 | 0.0444 | 0.008 | 2.58E-08 | Yes | Yes |
| rs35158675 | 1 | 222829550 | G | A | 0.6821 | 0.0699 | 0.0064 | 5.91E-28 | Yes | Yes |
| rs16986953 | 2 | 19942473 | G | A | 0.1095 | 0.0812 | 0.0098 | 1.06E-16 | Yes | Yes |
| rs515135 | 2 | 21286057 | C | T | 0.1837 | -0.0555 | 0.0066 | 5.74E-17 | Yes | Yes |
| rs6544713 | 2 | 44073881 | C | T | 0.3038 | 0.0492 | 0.0056 | 1.84E-18 | Yes | Yes |
| rs582384 | 2 | 45896437 | C | A | 0.5251 | 0.0332 | 0.0058 | 7.64E-09 | Yes | Yes |
| rs6743030 | 2 | 85763520 | C | T | 0.465 | 0.0565 | 0.0057 | 1.81E-23 | Yes | Yes |
| rs2252641 | 2 | 145801461 | C | T | 0.5299 | -0.0368 | 0.0051 | 5.41E-13 | Yes | Yes |
| rs12999907 | 2 | 164957251 | G | A | 0.8189 | 0.0482 | 0.0074 | 6.31E-11 | Yes | Yes |
| rs6728861 | 2 | 203873743 | G | A | 0.1174 | 0.1054 | 0.0089 | 1.29E-32 | Yes | Yes |
| rs1250229 | 2 | 216304384 | C | T | 0.2722 | 0.0435 | 0.0059 | 1.58E-13 | Yes | Yes |
| rs2571445 | 2 | 218683154 | G | A | 0.3895 | 0.037 | 0.0052 | 1.56E-12 | Yes | Yes |
| rs11677932 | 2 | 238223955 | G | A | 0.3157 | -0.0339 | 0.0061 | 2.64E-08 | Yes | Yes |
| rs34991912 | 3 | 14926351 | C | T | 0.4344 | 0.0379 | 0.0057 | 3.21E-11 | Yes | Yes |
| rs7633770 | 3 | 46688562 | G | A | 0.4061 | 0.0295 | 0.0052 | 1.10E-08 | Yes | Yes |
| rs7617773 | 3 | 48193515 | C | T | 0.6693 | 0.0355 | 0.0054 | 3.48E-11 | Yes | Yes |
| rs4678145 | 3 | 124450081 | G | C | 0.1284 | 0.0639 | 0.0083 | 1.45E-14 | Yes | Yes |
| rs10512861 | 3 | 132257961 | G | T | 0.1433 | -0.0431 | 0.0073 | 3.00E-09 | Yes | Yes |
| rs667920 | 3 | 136069472 | G | T | 0.7751 | 0.0489 | 0.0063 | 6.08E-15 | Yes | Yes |
| rs185244 | 3 | 138092889 | C | T | 0.1625 | 0.0644 | 0.0076 | 2.40E-17 | Yes | Yes |
| rs789294 | 3 | 154057741 | G | C | 0.1649 | -0.0642 | 0.0081 | 3.36E-15 | Yes | Yes |
| rs4266144 | 3 | 156852592 | G | C | 0.6774 | -0.0346 | 0.0061 | 1.40E-08 | Yes | Yes |
| rs12897 | 3 | 172115902 | G | A | 0.5914 | -0.0358 | 0.0059 | 1.21E-09 | Yes | Yes |
| rs17081933 | 4 | 57822869 | T | A | 0.1979 | 0.0399 | 0.007 | 1.17E-08 | Yes | Yes |
| rs12500824 | 4 | 77416627 | G | A | 0.3565 | 0.0336 | 0.0053 | 1.64E-10 | Yes | Yes |
| rs10857147 | 4 | 81181072 | T | A | 0.7208 | -0.0491 | 0.0064 | 1.37E-14 | Yes | Yes |
| rs11099493 | 4 | 82587050 | G | A | 0.6924 | 0.039 | 0.0062 | 2.49E-10 | Yes | Yes |
| rs3775058 | 4 | 96117371 | T | A | 0.2341 | 0.0386 | 0.0067 | 7.63E-09 | Yes | Yes |
| rs7678555 | 4 | 120909501 | C | A | 0.7166 | -0.0486 | 0.0063 | 1.23E-14 | Yes | Yes |
| rs6841581 | 4 | 148401190 | G | A | 0.1559 | 0.0751 | 0.0074 | 5.18E-24 | Yes | Yes |
| rs7692387 | 4 | 156635309 | G | A | 0.1892 | -0.0643 | 0.0065 | 3.34E-23 | Yes | Yes |
| rs7696431 | 4 | 169687725 | G | T | 0.5098 | 0.0311 | 0.0056 | 3.09E-08 | Yes | Yes |
| rs112941079 | 5 | 9546098 | G | A | 0.8676 | 0.0615 | 0.0085 | 3.75E-13 | Yes | Yes |
| rs3936511 | 5 | 55860781 | G | A | 0.823 | -0.0366 | 0.0067 | 3.74E-08 | Yes | Yes |
| rs273909 | 5 | 131667353 | G | A | 0.8754 | -0.0488 | 0.0078 | 4.74E-10 | No | Yes |
| rs246600 | 5 | 142516897 | C | T | 0.4633 | 0.043 | 0.0051 | 6.53E-17 | Yes | Yes |
| rs9349379 | 6 | 12903957 | G | A | 0.5907 | -0.1072 | 0.0058 | 2.72E-76 | Yes | Yes |
| rs6909752 | 6 | 22612629 | G | A | 0.3545 | 0.0442 | 0.0059 | 5.71E-14 | Yes | Yes |
| rs644045 | 6 | 31883957 | G | A | 0.3528 | -0.0413 | 0.0057 | 2.80E-13 | Yes | No |
| rs2814944 | 6 | 34552797 | G | A | 0.1554 | 0.0479 | 0.0072 | 2.15E-11 | Yes | Yes |
| rs1334894 | 6 | 35615130 | C | T | 0.0919 | -0.0482 | 0.0088 | 4.84E-08 | Yes | Yes |
| rs1321309 | 6 | 36638636 | G | A | 0.4907 | 0.028 | 0.0051 | 3.48E-08 | Yes | Yes |
| rs10947789 | 6 | 39174922 | C | T | 0.7671 | 0.0413 | 0.006 | 7.63E-12 | Yes | Yes |
| rs6905288 | 6 | 43758873 | G | A | 0.5807 | 0.0386 | 0.0056 | 4.00E-12 | Yes | Yes |
| rs71566846 | 6 | 57148171 | C | T | 0.0662 | 0.0829 | 0.0114 | 3.24E-13 | Yes | Yes |
| rs4613862 | 6 | 82612271 | C | A | 0.5314 | 0.0319 | 0.0052 | 6.58E-10 | Yes | Yes |
| rs1591805 | 6 | 126717064 | G | A | 0.4927 | 0.037 | 0.0058 | 1.17E-10 | Yes | Yes |
| rs2327429 | 6 | 134209837 | C | T | 0.695 | 0.0655 | 0.0056 | 3.56E-31 | Yes | Yes |
| rs17080091 | 6 | 150997401 | C | T | 0.0839 | -0.0537 | 0.0092 | 6.09E-09 | Yes | Yes |
| rs55730499 | 6 | 161005610 | C | T | 0.0693 | 0.3122 | 0.0118 | 9.78E-154 | Yes | Yes |
| rs10267593 | 7 | 1937261 | G | A | 0.2012 | -0.036 | 0.0064 | 1.88E-08 | Yes | Yes |
| rs7797644 | 7 | 6486067 | C | T | 0.2321 | -0.0386 | 0.0069 | 2.10E-08 | Yes | Yes |
| rs11509880 | 7 | 12261911 | G | A | 0.3612 | 0.0327 | 0.0059 | 3.27E-08 | Yes | Yes |
| rs2107595 | 7 | 19049388 | G | A | 0.1767 | 0.0752 | 0.0073 | 1.25E-24 | Yes | Yes |
| rs2107732 | 7 | 45077978 | G | A | 0.0874 | -0.0567 | 0.0103 | 3.64E-08 | No | Yes |
| rs2189839 | 7 | 107230026 | G | A | 0.2899 | 0.0345 | 0.0063 | 4.53E-08 | Yes | Yes |
| rs975722 | 7 | 117332914 | G | A | 0.595 | -0.0283 | 0.0052 | 4.17E-08 | Yes | Yes |
| rs11556924 | 7 | 129663496 | C | T | 0.3624 | -0.0548 | 0.0055 | 1.37E-23 | Yes | Yes |
| rs10237377 | 7 | 139757136 | G | T | 0.3608 | -0.0338 | 0.0058 | 6.53E-09 | Yes | Yes |
| rs3918226 | 7 | 150690176 | C | T | 0.0732 | 0.1071 | 0.0115 | 1.35E-20 | Yes | Yes |
| rs6997340 | 8 | 18286997 | C | T | 0.3025 | 0.0331 | 0.0056 | 4.60E-09 | Yes | Yes |
| rs17091891 | 8 | 19843171 | C | T | 0.8709 | 0.0585 | 0.0078 | 5.15E-14 | Yes | Yes |
| rs6984210 | 8 | 22033615 | G | C | 0.939 | -0.0784 | 0.0115 | 1.04E-11 | No | Yes |
| rs10093110 | 8 | 106565414 | G | A | 0.4167 | -0.0319 | 0.0057 | 1.88E-08 | Yes | Yes |
| rs6982502 | 8 | 126479362 | C | T | 0.5298 | -0.0498 | 0.0051 | 7.67E-23 | Yes | Yes |
| rs4977574 | 9 | 22098574 | G | A | 0.519 | -0.1788 | 0.0056 | 8.82E-223 | Yes | Yes |
| rs944172 | 9 | 110517794 | C | T | 0.7197 | -0.0395 | 0.0057 | 3.59E-12 | Yes | Yes |
| rs885150 | 9 | 124420173 | C | T | 0.7304 | -0.0355 | 0.0058 | 7.86E-10 | Yes | Yes |
| rs2519093 | 9 | 136141870 | C | T | 0.1841 | 0.0554 | 0.0072 | 2.03E-14 | Yes | Yes |
| rs61848342 | 10 | 12303813 | C | T | 0.6355 | -0.0363 | 0.0059 | 6.38E-10 | Yes | Yes |
| rs9337951 | 10 | 30317073 | G | A | 0.3178 | 0.0543 | 0.0064 | 1.73E-17 | No | Yes |
| rs1870634 | 10 | 44480811 | G | T | 0.3511 | -0.06 | 0.0059 | 3.50E-24 | Yes | Yes |
| rs17680741 | 10 | 82251514 | C | T | 0.7152 | 0.042 | 0.0062 | 1.72E-11 | Yes | Yes |
| rs1412444 | 10 | 91002927 | C | T | 0.3527 | 0.0559 | 0.0059 | 2.43E-21 | Yes | Yes |
| rs3740390 | 10 | 104638480 | C | T | 0.1066 | -0.0662 | 0.0084 | 4.67E-15 | Yes | Yes |
| rs4918072 | 10 | 105693644 | G | A | 0.2713 | 0.0386 | 0.0063 | 9.63E-10 | Yes | Yes |
| rs4752700 | 10 | 124237612 | G | A | 0.5508 | -0.0332 | 0.0051 | 8.02E-11 | Yes | Yes |
| rs11601507 | 11 | 5701074 | C | A | 0.0739 | 0.0782 | 0.0108 | 5.61E-13 | Yes | Yes |
| rs472109 | 11 | 9770318 | G | C | 0.5698 | 0.0391 | 0.0057 | 6.26E-12 | Yes | Yes |
| rs7926712 | 11 | 13303085 | G | A | 0.3101 | -0.0351 | 0.0061 | 9.41E-09 | Yes | Yes |
| rs7116641 | 11 | 43696917 | G | T | 0.6877 | -0.0314 | 0.0055 | 1.03E-08 | Yes | Yes |
| rs12801636 | 11 | 65391317 | G | A | 0.2364 | -0.0403 | 0.006 | 2.29E-11 | Yes | Yes |
| rs606452 | 11 | 75276178 | C | A | 0.1707 | -0.0466 | 0.0071 | 4.76E-11 | Yes | Yes |
| rs4754698 | 11 | 100631908 | G | C | 0.5399 | -0.0368 | 0.0056 | 6.79E-11 | Yes | Yes |
| rs974819 | 11 | 103660567 | C | T | 0.3082 | 0.0614 | 0.0055 | 1.12E-28 | Yes | Yes |
| rs651821 | 11 | 116662579 | C | T | 0.901 | -0.0692 | 0.0096 | 7.03E-13 | Yes | Yes |
| rs11838267 | 12 | 7175872 | C | T | 0.8679 | 0.0514 | 0.0083 | 6.15E-10 | No | Yes |
| rs10841443 | 12 | 20220033 | G | C | 0.3409 | -0.046 | 0.0061 | 2.86E-14 | Yes | Yes |
| rs11170820 | 12 | 54513915 | G | C | 0.9292 | -0.0832 | 0.0117 | 9.28E-13 | Yes | Yes |
| rs11613352 | 12 | 57792580 | C | T | 0.235 | -0.036 | 0.0062 | 4.92E-09 | Yes | Yes |
| rs2681492 | 12 | 90013089 | C | T | 0.8141 | -0.0558 | 0.0072 | 8.64E-15 | Yes | Yes |
| rs11107903 | 12 | 95507971 | G | A | 0.0768 | -0.0747 | 0.0105 | 1.06E-12 | Yes | Yes |
| rs7137828 | 12 | 111932800 | C | T | 0.5535 | -0.0643 | 0.006 | 3.72E-27 | Yes | Yes |
| rs1169288 | 12 | 121416650 | C | A | 0.6725 | -0.049 | 0.0056 | 1.26E-18 | Yes | Yes |
| rs11057830 | 12 | 125307053 | G | A | 0.1465 | 0.0655 | 0.0075 | 1.91E-18 | Yes | Yes |
| rs9319428 | 13 | 28973621 | G | A | 0.3121 | 0.036 | 0.0055 | 5.36E-11 | Yes | Yes |
| rs7998440 | 13 | 33125206 | G | A | 0.3549 | -0.0383 | 0.0059 | 1.19E-10 | Yes | Yes |
| rs9515203 | 13 | 111049623 | C | T | 0.7392 | 0.0596 | 0.006 | 3.89E-23 | No | Yes |
| rs1317507 | 13 | 113631780 | C | A | 0.2608 | 0.0398 | 0.0058 | 8.21E-12 | Yes | Yes |
| rs2145598 | 14 | 58794001 | G | A | 0.5758 | -0.0283 | 0.0052 | 4.26E-08 | Yes | Yes |
| rs7145159 | 14 | 75583268 | C | T | 0.5234 | -0.0319 | 0.0051 | 3.60E-10 | Yes | Yes |
| rs112635299 | 14 | 94838142 | G | T | 0.0193 | -0.1363 | 0.0222 | 8.44E-10 | Yes | Yes |
| rs8003602 | 14 | 100148961 | C | T | 0.2681 | -0.0539 | 0.0066 | 2.89E-16 | Yes | Yes |
| rs6494488 | 15 | 65024204 | G | A | 0.8117 | 0.0382 | 0.007 | 3.90E-08 | Yes | Yes |
| rs72743461 | 15 | 67441750 | C | A | 0.2211 | -0.0576 | 0.0069 | 5.68E-17 | Yes | Yes |
| rs7173743 | 15 | 79141784 | C | T | 0.5542 | 0.0636 | 0.0051 | 5.48E-36 | Yes | Yes |
| rs1807214 | 15 | 89565257 | C | A | 0.8891 | 0.0639 | 0.0097 | 5.21E-11 | Yes | Yes |
| rs17514846 | 15 | 91416550 | C | A | 0.4605 | 0.0559 | 0.0052 | 9.86E-27 | Yes | Yes |
| rs17581137 | 15 | 96146414 | C | A | 0.754 | 0.0373 | 0.0065 | 1.21E-08 | Yes | Yes |
| rs12149545 | 16 | 56993161 | G | A | 0.3035 | -0.0374 | 0.0062 | 1.19E-09 | Yes | Yes |
| rs1050362 | 16 | 72130815 | C | A | 0.3755 | 0.0352 | 0.0053 | 2.92E-11 | Yes | Yes |
| rs8046696 | 16 | 75442143 | G | T | 0.4311 | -0.0479 | 0.0058 | 1.91E-16 | Yes | Yes |
| rs7199941 | 16 | 81906423 | G | A | 0.3982 | 0.0367 | 0.0051 | 9.36E-13 | Yes | Yes |
| rs7500448 | 16 | 83045790 | G | A | 0.7594 | 0.0557 | 0.0068 | 1.61E-16 | Yes | Yes |
| rs170041 | 17 | 2170216 | C | T | 0.2943 | -0.0469 | 0.0056 | 4.11E-17 | Yes | Yes |
| rs12936587 | 17 | 17543722 | G | A | 0.4379 | -0.032 | 0.0052 | 9.51E-10 | Yes | Yes |
| rs11080107 | 17 | 27938424 | C | T | 0.5146 | -0.0358 | 0.0057 | 2.90E-10 | Yes | Yes |
| rs76954792 | 17 | 30033514 | C | T | 0.2197 | 0.0393 | 0.0069 | 1.19E-08 | Yes | Yes |
| rs2074164 | 17 | 40270238 | G | C | 0.1956 | 0.0423 | 0.0073 | 7.36E-09 | Yes | Yes |
| rs17608766 | 17 | 45013271 | C | T | 0.8632 | -0.0469 | 0.0076 | 8.20E-10 | Yes | No |
| rs62076439 | 17 | 47404628 | G | T | 0.3409 | 0.048 | 0.006 | 1.63E-15 | Yes | Yes |
| rs1476098 | 17 | 59237013 | C | A | 0.2122 | 0.0443 | 0.0072 | 8.51E-10 | Yes | Yes |
| rs9892152 | 17 | 62401965 | C | T | 0.4717 | -0.033 | 0.005 | 6.28E-11 | Yes | Yes |
| rs9964304 | 18 | 47229717 | C | A | 0.716 | -0.0382 | 0.0063 | 1.14E-09 | Yes | Yes |
| rs663640 | 18 | 57846077 | C | T | 0.222 | 0.0383 | 0.0068 | 2.02E-08 | Yes | Yes |
| rs116843064 | 19 | 8429323 | G | A | 0.0202 | -0.1402 | 0.0224 | 3.57E-10 | Yes | Yes |
| rs55791371 | 19 | 11188153 | C | A | 0.8868 | 0.1157 | 0.0092 | 1.93E-36 | Yes | Yes |
| rs7251815 | 19 | 17844942 | G | T | 0.2162 | 0.0511 | 0.0069 | 1.44E-13 | Yes | Yes |
| rs4803455 | 19 | 41851509 | C | A | 0.4954 | -0.0484 | 0.0057 | 2.36E-17 | Yes | Yes |
| rs7412 | 19 | 45412079 | C | T | 0.079 | -0.1368 | 0.011 | 2.14E-35 | Yes | Yes |
| rs867186 | 20 | 33764554 | G | A | 0.8971 | 0.0573 | 0.0084 | 6.84E-12 | Yes | Yes |
| rs6102343 | 20 | 39924279 | G | A | 0.2467 | 0.0372 | 0.0065 | 1.12E-08 | Yes | Yes |
| rs3827066 | 20 | 44586023 | C | T | 0.1437 | 0.0424 | 0.0072 | 4.40E-09 | Yes | Yes |
| rs260020 | 20 | 57714025 | C | T | 0.1309 | 0.0518 | 0.0084 | 7.96E-10 | Yes | Yes |
| rs2832227 | 21 | 30533076 | G | A | 0.8258 | -0.0393 | 0.0067 | 4.16E-09 | Yes | Yes |
| rs28451064 | 21 | 35593827 | G | A | 0.1239 | 0.1083 | 0.009 | 2.58E-33 | Yes | Yes |

**Supplemental Table 2.** Sample sizes.

|  | Women |  |  | Male partners |  |  |
| --- | --- | --- | --- | --- | --- | --- |
|  | Main<br>analyses | Only stable<br>couples | Both parents with<br>genotype data | Main<br>analyses | Only stable<br>couples | Both parents with<br>genotype data |
| Miscarriage | 75,210 | 65,590 | 45,273 | 51,156 | 45,239 | 45,212 |
| Stillbirth | 83,900 | 74,009 | 51,396 | 55,790 | 49,794 | 49,846 |
| Hypertensive disorders of pregnancy | 83,114 | 73,339 | 50,906 | 55,273 | 49,341 | 49,377 |
| Pre-eclampsia + eclampsia | 83,114 | 73,339 | 50,906 | 55,273 | 49,341 | 49,377 |
| Gestational diabetes | 83,900 | 74,009 | 51,396 | 55,790 | 49,794 | 49,846 |
| Small for gestational age | 66,110 | 58,857 | 40,703 | 43,967 | 39,395 | 39,334 |
| Large for gestational age | 69,789 | 61,994 | 42,972 | 46,104 | 41,324 | 41,293 |
| Spontaneous preterm birth | 83,522 | 73,640 | 51,167 | 55,536 | 49,595 | 49,621 |

**Supplemental Table 3.** Description of participants with and without genotype data in MoBa and HUNT.

|  | MoBa |  |  |  | HUNT |  |  |  |
| --- | --- | --- | --- | --- | --- | --- | --- | --- |
|  | Women |  | Men |  | Women |  | Men |  |
|  | Included | Non-included | Included | Non-included | Included | Non-included | Included | Non-included |
| Birth year,<br>median (1<sup>st</sup>-3<sup>rd</sup> quartile) | 1975<br>(1972-1979) | 1975<br>(1972-1979) | 1973<br>(1970-1977) | 1973<br>(1969-1976) | 1964<br>(1958-1971) | 1976<br>(1966-1985) | 1962<br>(1956-1969) | 1974<br>(1964-1983) |
| Number of pregnancies,<br>median (1<sup>st</sup>-3<sup>rd</sup> quartile) | 2<br>(2-3) | 2<br>(2-3) | 2<br>(2-3) | 2<br>(2-3) | 2<br>(2-3) | 2<br>(2-3) | 2<br>(2-3) | 2<br>(2-3) |
| Participants in pregnancies<br>with more than one different<br>partner, *n* (%) | 9,147<br>(13.3%) | 2,099<br>(13.1%) | 5,819<br>(12.3%) | 2,945<br>(13.9%) | 745<br>(4.74%) | 203<br>(3.53%) | 178<br>(2.08%) | 51<br>(1.70%) |
| Age in first pregnancy,<br>median (1<sup>st</sup>-3<sup>rd</sup> quartile) | 27<br>(24-30) | 27<br>(24-30) | 29<br>(26-33) | 29<br>(26-33) | 24<br>(20-27) | 24<br>(21-28) | 26<br>(23-29) | 27<br>(24-30) |
| Education years, mean ± SD | 17.0 ± 3.36 | 16.7 ± 3.55 | 16.3 ± 3.56 | 16.0 ± 3.66 | 14.3 ± 4.13 | 15.0 ± 4.14 | 13.5 ± 4.08 | 14.1 ± 4.27 |
| Body mass index (kg/m<sup>2</sup>), | 23.1 | 23.0 | 25.4 | 25.4 | 25.2 | 25.3 | 26.6 | 26.7 |
| median (1<sup>st</sup>-3<sup>rd</sup> quartile) | (21.1-25.9) | (20.9-25.7) | (23.6-27.7) | (23.6-27.7) | (22.7-28.6) | (22.6-29.0) | (24.5-29.1) | (24.5-29.4) |
| Ever smokers, *n* (%) | 35,632<br>(52.3%) | 8,071<br>(51.1%) | 24,294<br>(51.2%) | 11,461<br>(54.1%) | 8,865<br>(57.0%) | 3,080<br>(54.0%) | 4,233<br>(50.1%) | 1,533<br>(51.4%) |
| Total number of deliveries,<br>mean ± SD | 2.54 ± 0.85 | 2.55 ± 0.89 | 2.51 ± 0.81 | 2.54 ± 0.86 | 1.48 ± 0.88 | 1.28 ± 0.94 | 1.50 ± 0.81 | 1.31 ± 0.85 |
| Miscarriage, *n* (%) | 20,866<br>(30.4%) | 4,835<br>(30.7%) | 14,451<br>(30.6%) | 6,484<br>(31.0%) | 1,950<br>(30.0%)<sup>a</sup> | 1,184<br>(29.4%)<sup>a</sup> | 1,145<br>(29.6%)<sup>a</sup> | 662<br>(29.3%)<sup>a</sup> |
| Stillbirth, *n* (%) | 872<br>(1.27%) | 238<br>(1.51%) | 581<br>(1.23%) | 313<br>(1.49%) | 198<br>(1.30%) | 45<br>(0.84%) | 84<br>(0.99%) | 19<br>(0.64%) |
| Any hypertensive disorders<br>of pregnancy, *n* (%) | 7,288<br>(10.6%) | 1,730<br>(11.0%) | 5,186<br>(11.0%) | 2,297<br>(11.0%) | 1,500<br>(9.87%) | 483<br>(9.02%) | 870<br>(10.2%) | 288<br>(9.66%) |
| Pre-eclampsia + eclampsia,<br>*n* (%) | 5,143<br>(7.49%) | 1,198<br>(7.61%) | 3,589<br>(7.59%) | 1,606<br>(7.67%) | 1,080<br>(7.11%) | 336<br>(6.28%) | 641<br>(7.54%) | 205<br>(6.88%) |
| Gestational diabetes, *n* (%) | 1,495<br>(2.18%) | 474<br>(3.01%) | 1,041<br>(2.20%) | 553<br>(2.64%) | 127<br>(0.84%) | 111<br>(2.07%) | 62<br>(0.73%) | 69<br>(2.31%) |
| Small for gestational age, | 10,552 | 2,706 | 7,354 | 3,608 | 1,544 | 251 | 862 | 136 |
| *n* (%) | (15.8%) | (17.7%) | (16.0%) | (17.8%) | (10.2%) | (4.69%) | (10.1%) | (4.56%) |
| Large for gestational age, | 14,620 | 3,063 | 9,880 | 4,375 | 1,733 | 285 | 1,070 | 179 |
| *n* (%) | (21.7%) | (20.0%) | (21.3%) | (21.4%) | (11.4%) | (5.33%) | (12.6%) | (6.00%) |
| Spontaneous preterm birth, | 6,282 | 1,508 | 4,187 | 2,027 | 2,643 | 688 | 1,509 | 385 |
| *n* (%) | (9.18%) | (9.63%) | (8.89%) | (9.73%) | (17.5%) | (13.0%) | (17.8%) | (13.0%) |

<sup>a</sup>: analyses restricted to 1998 or later pregnancies.

## SUPPLEMENTAL FIGURES

**Supplemental Figure 1.**
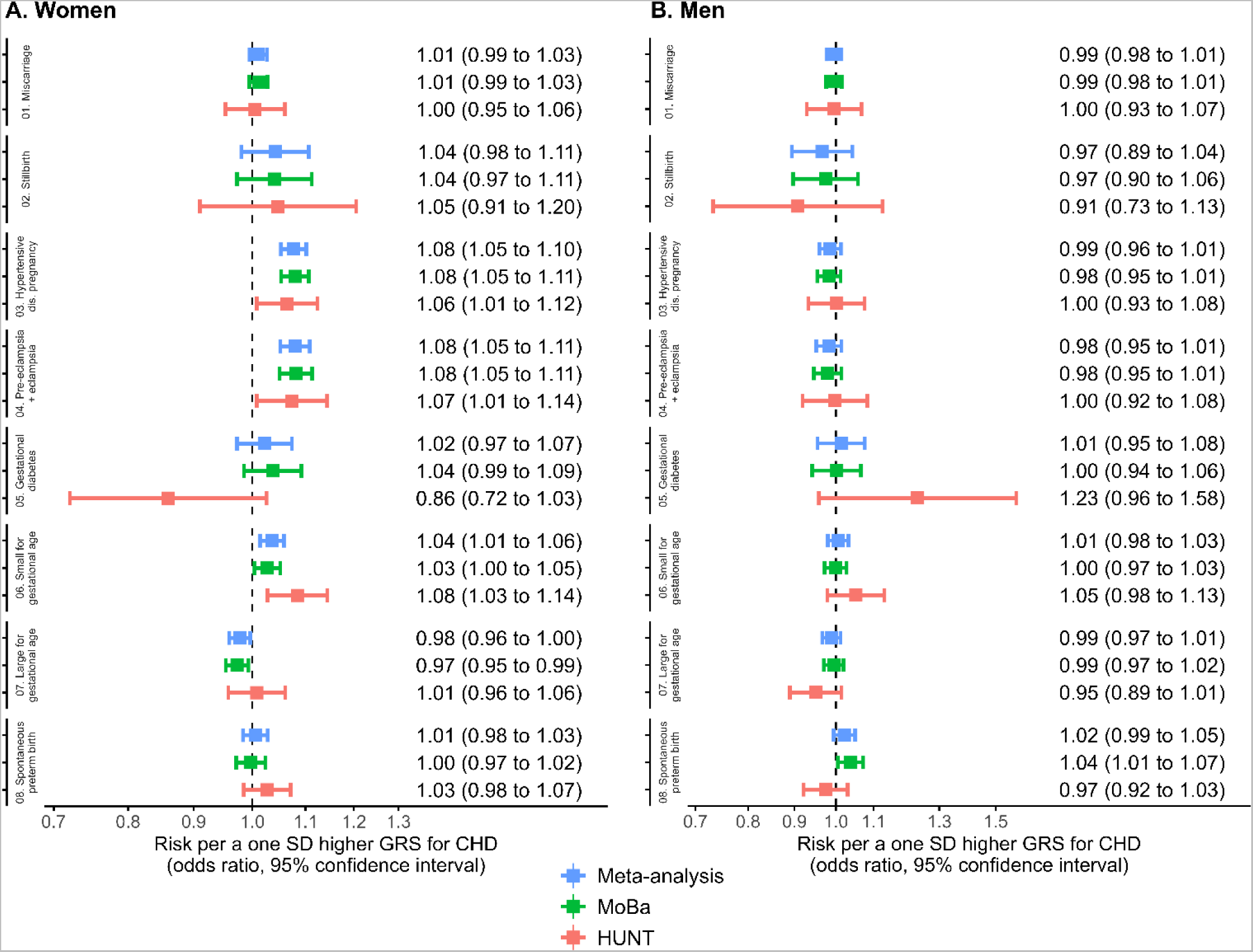
Associations between 1 SD increase in the genetic risk score for coronary heart disease and pregnancy outcomes in women (A) and men (B) in MoBa and HUNT participants individually. Analyses are adjusted for the first 20 ancestry-informative genetic principal components and genotype batch.

